# The Knowledge, attitude and practices about bio-medical waste management system in a tertiary care 60 bedded hospital in Rajasthan, India

**DOI:** 10.1101/2020.06.21.20136796

**Authors:** Virender Verma, Priya Soni, Meenakshi Kalhan, Sanjiv Nanda, Anjani Kumar, Vivek Nandan

## Abstract

**Background and Purpose:** From antiquity, the healthcare facilities have treated sick people but there was little awareness about the fact that hospitals generate a lot of hazardous bio-medical waste, which had been disposed without proper guidelines. Lately, it has been a proven thing that bio-medical-waste is a potential hazard for heath-personals, and environment. Therefore, it must be disposed in a proper manner.

**Objectives:** This was an observational study carried out at one of the tertiary care hospitals in Rajgarh city about the knowledge, attitude and practices of the hospital staff about the bio-medical waste management.

**Material and method:** An observational (cross-sectional) study using questionnaires was carried out at one of the non-NABH accredited tertiary care hospitals of Rajgarh City in Rajasthan (India).

**Results:** More than 70% staff (except housekeeping) had good ideas about waste categories, segregation and color coding except in radioactive waste. The house keeping staff did well in 3 categories (Linen, sharps and glasses). Most of the employees has clarity about BMW except the housekeeping staff which did well on most of parameters except barcoding, pretreatment of anatomical/ biotechnology waste, knowledge about STP plant (Sewage treatment plant), Hazmat and signage. On actual hospital rounds we found that the hazmat practices were in a poor shape. Most of staff was aware occupational hazards except housekeeping staff.

**Conclusion:** A written policy, induction training of healthcare workers, constant and repetitive workshops, and motivation are important human factors to implement the biomedical waste practices in a small-scale healthcare organization. Risk-stratification, understanding of health hazards and how to activate hazmat protocols are important things which are to be brought into practice.

## Introduction

Human activities and interventions generate some useful products and some by-products; these generally useful by-products are often termed as waste. It has been a proven thing that bio-medical-waste is an important potential hazard for heath-personals, and environment [1]. Previously when human population was less, the waste generated by human actives was taken care of by natural phenomenon like bacterial degradation, saprophytes and natural scavengers, but the second half of 20^th^ century witnessed an exponential rise in human activities resulting into enormous waste whose management has resulted into a formulation of many guidelines and practices [2].

For the purpose of definition [3]:

“**Hospital waste** means collective waste, (biological or non-biological) which is not useful and discarded”

**“Bio**-**medical waste** refers to waste generated during the diagnosis, treatment or immunization of human beings or animals or in research activities or in the production or testing of biologicals.”

**“Infectious waste:** The by-products which contain or might contain pathogens in sufficient concentration that could cause diseases for example, culture of infectious agents from laboratories, waste from operation theatres, or from infectious patients.”

An act was passed by the Ministry of Environment and Forests in 1986 & notified in July 1998, “**The Bio Medical Waste (Management and Handling) Rules”**, which states that, “Ensuring disposal of waste generated by an institution in a proper way so that it leaves no hazardous effect on human health can environment, is the duty of every occupant of the institution and vicariously reflected by the head [4].

Despite many laws and reinforcements by state and central agencies the bio-medical waste is still a difficult practice at ground level, imposing a considerable health risk on health care workers, patients and environment [1]. Bio-medical-waste forms 1-2% of total solid waste collection by municipality in general [5]. In India, an average hospital bed generates around two kilograms of bio-waste per day [5]. As alluded earlier, BMW might be full of disease causing organisms viz hepatitis B, hepatitis C, and AIDS [1]. Therefore, its proper handling is vital. The BMW management practices are a collective effort of higher administration (policy and funding), biomedical engineering (technology and equipment) and Operation / housekeeping department (training and induction), immediate healthcare workers (doctors, nurses and paramedics) and ground force of housekeeping department [6, 7].

The Biomedical waste 2004 guidelines of WHO emphasize the important of “Human Factor” over technology and equipment. Staff motivation and dedication is vital for success of proper disposal of BMW, highly motivated staff can compensate limitation arising because of lack of sophisticated equipment in small health care organizations because of limited budget [8]. According to a government statement there are a total 37725 health-care facilities and approximately 3500 Ayush centers were operational in India in 2018 [9]. There are a lot of private hospitals which are unregistered and producing BMW. We conducted this study in a small health care organization (60 bedded) at Rajgarh city (Rajasthan) in India.

### Ethical Consideration

Ethical approval was taken form the institutional ethical committee and all the measures were taken to ensure Helsinki declaration. Informed and written consent to participate in the study was obtained by the participants.

## Materials and Methods

### Subjects

#### Inclusion Criteria

We included all the hospital employees regardless of their hierarchy.

#### Exclusion Criteria

We did not include trainees, security staff and marketing ground force as formers were outsourced and latter were hardly having any function inside the hospital premises and BMW generation and disposal directly or indirectly.

### Methodology

We conducted this observational (cross section) study at a tertiary care small health care organization (SHCO), which has recently considered for applying to nation accreditation board of hospital (NABH), an accreditation agency for hospital quality in India. The hospital is a 60 bedded multispecialty allopathic hospital with all major surgical and medical specialties. It has a routine OPD of almost 250 patients and in-house occupancy rates averaging 80%. The hospital has around 8 higher management officials, 1 quality manager, 4 other managers, 1 bio-medical engineer, 4 pharmacists, 17 doctors, 52 nurses, 4 Lab-technologists and 74 other support staff (housekeeping staff). We carried out a survey about knowledge attitude and practices about BMW using a questionnaire.

A scientifically designed questionnaire, which included details about knowledge of infectivity, disposal (segregation, transport and final disposal), knowledge and attitude about immunization and health hazards and use of protective equipment in the management of BMW (Bio-medical-waste) was used. We did not include personal details and duration of the person in healthcare organization in order to get the unbiased and honest answers. However, the questionnaires were printed on colored papers for subgroup analysis, that is, white paper for higher management, managers and pharmacists, pink for nurses and lab-technologists, orange for doctors and blue for housekeeping staff. These were printed in English as well as Local language (Hindi), and handed over to respected staff by the human resource manager in opaque envelope. At the time of handing over the employee was thoroughly explained about the need of honesty and it was reinforced that this will be used for training purpose and must not affect their employability. These were distributed between 1^st^ to 5^th^ of January 2020 and the hospital employees were directed to return them without fail on or before 10^th^ of the January in the office of Human resource manager, they day they come to receive their salary cheques. This was step was taken to maximize responses. When the person came for receiving cheque, they needed to drop the envelope containing the response in an opaque letter box and tick on a checklist containing their name. Those employees who did not return the questionnaire by 10^th^ were contacted on their mobiles by Human resource manager and were asked to return it at earliest. All questionnaires were received by 13^th^ of the January 2020. The box was shaken and opened in presence of principal investigator and all questionnaires were taken out. These were segregated using the color codes.

Out of 165 responses 6 were rejected because of difficult to understand handwriting, ambiguous responses which did not make any sense or blank response etc. 159 questionnaires were analyzed, however, the total number for calculation was assumed to be 165 and same as initial values in individual categories for the purpose of calculations. On 24^th^ and 25^th^ of January hospital rounds were taken to observe and record the real practices about BMW management and to know the belief of the employees right at the point of care.

## Results

Table 1 depicts the actual number (percentage) of employees their gender and age and educational qualification. It is observed that 87% nurses and Lab technicians and 35% housekeeping staff were females. Majority of employees were in the age group of 25years to 40 years. Most of nurses were diploma holder (76%) or higher (24%), all doctors were graduate and above and only 11% house-keeping staff was educated beyond school.

**Table 1:**
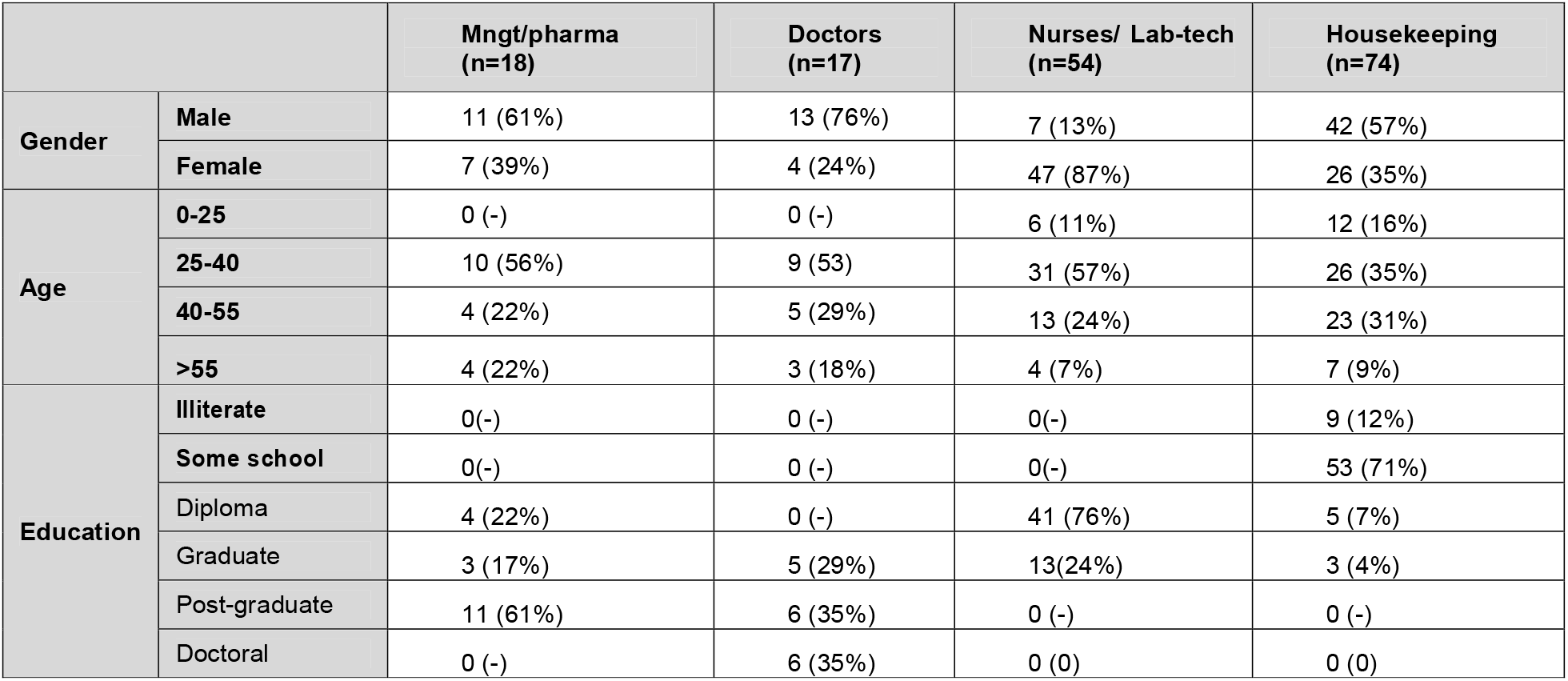
Demographic and educational details of study-subjects.

The employee composition is primarily a young and middle age group type.

Table 2 depicts level of understanding of various categories of waste material and their segregation into various bins/boxes. It is seen that most of management person, doctors and nurses did well in most of categories and more than 70% had good ideas about it except in radioactive waste where they scored poor around 30%. The house keeping staff did well in 3 categories (Linen, sharps and glasses) and scored about 70% correct responses.

**Table 2:**
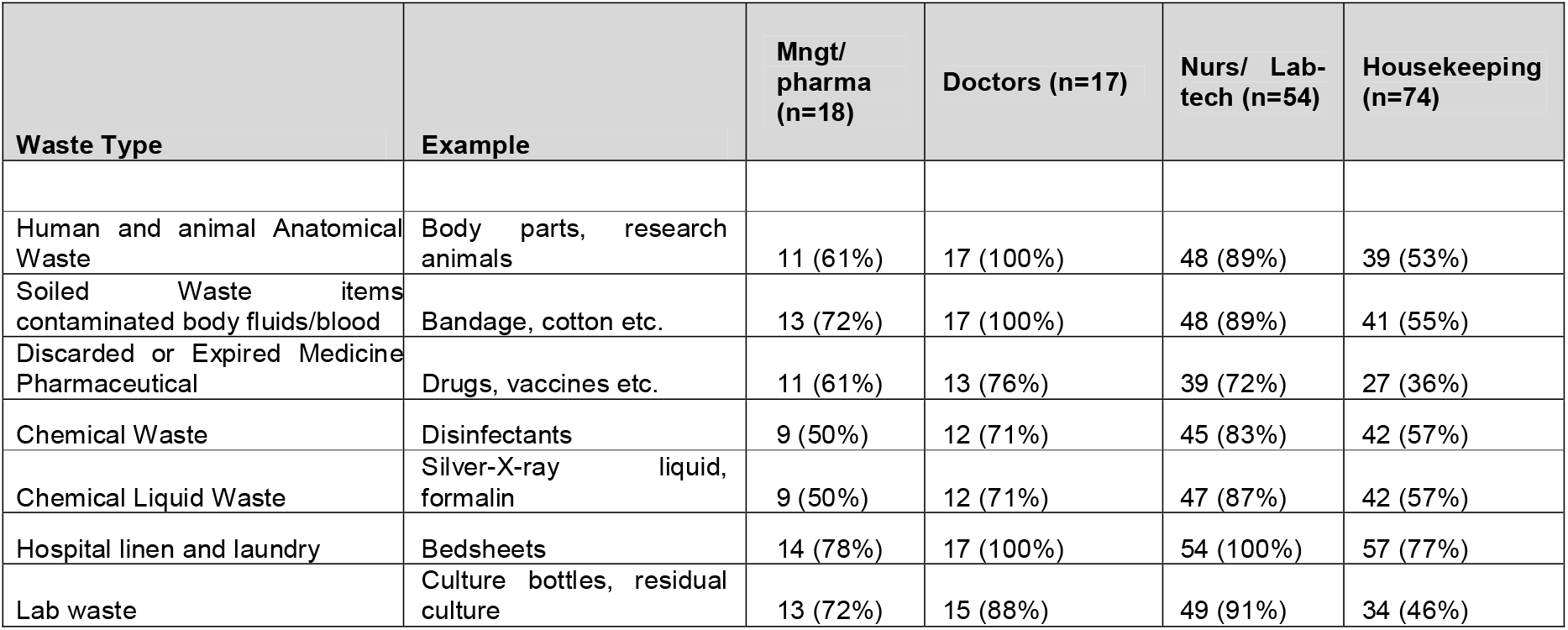
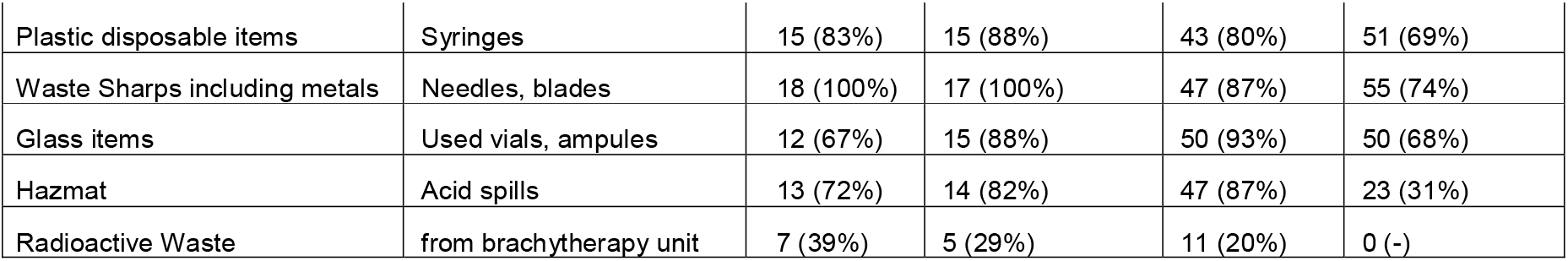
Knowledge of employees about waste categories and color coding etc.

In Table 3, the operation and quality measures have been depicted and it is clear that most of employees has a clear idea about BMW operation and quality measures except the housekeeping staff which did well on most of parameters except barcoding, pretreatment of anatomical and biotechnology waste, knowledge about STP (Sewage-treatment-plant) [10], Hazmat [11, 12] and signage. On actual hospital rounds we found that the hazmat practices were in a poor shape when even nurses could not brief about them. The barcode system was not followed yet although managers, doctors and nurses had dependable knowledge.

**Table 3:** BMW practices: operations and quality knowledge and actual observed compliance

| Operation and quality | Mngt/pharma<br>(n=18) | Doctors<br>(n=17) | Nurses/<br>Lab-tech<br>(n=54) | Housekeeping<br>(n=74) | Observed Compliance |  |  |
| --- | --- | --- | --- | --- | --- | --- | --- |
|  |  |  |  |  | Ward | Lab | OT |
| Segregation at site of generation | 18 (100%) | 17 (100%) | 54 (100%) | 68 (92%) | 100% | 100% | 100% |
| Color coding of bags and container | 18 (100%) | 17 (100%) | 54 (100%) | 68 (92%) | 100% | 100% | 100% |
| Labelling of containers | 16 (89%) | 17 (100%) | 54 (100%) | 68 (92%) | 100% | 100% | 100% |
| Bar code and global positioning | 14 (78%) | 13 (76%) | 36 (67%) | **10 (14%)** | - | - | - |
| Quality of transport to vehicle | 18 (100%) | 17 (100%) | 43 (80%) | 68 (92%) | 100% | 100% | 100% |
| Transport vehicle to facility | 18 (100%) | 17 (100%) | 40 (74%) | 68 (92%) | 100% | 100% | 100% |
| Pretreatment of anatomical, soiled waste etc. if stored | 18 (100%) | 15 (88%) | 39 (72%) | **27 (36%)** | NA | NA | NA |
| Treatment of microbiology waste | 18 (100%) | 17 (100%) | 52 (96%) | **10 (14%)** | - | 100% |  |
| STP plant | 18 (100%) | 16 (94%) | 40 (74%) | **10 (14%)** | + | + | + |
| Hazmat practices | 13 (72%) | 14 (82%) | 47 (87%) | **23 (31%)** | **32%** | **67%** | **50%** |
| Signage and charts | 16 (89%) | 15 (88%) | 36 (67%) | **18 (24%)** | + | + | + |
| Maintenance of record | 18 (100%) | 17 (100%) | 54 (100%) | 68 (92%) | + | + | + |
| Ample disinfectant available | 18 (100%) | 17 (100%) | 54 (100%) | 68 (92%) | + | + | + |

Table 4 shows that most of staff was aware of tetanus and hepatitis B vaccination except housekeeping staff (16% had some idea about hepatitis B). Thirty five percent housekeeping staff were knowing that they need to immediately inform the laboratory incharge in case of needle prick and need to get some tests and take medicines.

**Table 4:** KAP about occupational safety

| Occupational safety | Mngt/pharma<br>(n=18) | Doctors<br>(n=17) | Nurses/<br>Lab-tech<br>(n=54) | Housekeepin<br>g (n=74) | Observed Compliance |  |  |
| --- | --- | --- | --- | --- | --- | --- | --- |
|  |  |  |  |  | Ward | Lab | OT |
| TT immunization | 18 (100%) | 17 (100%) | 54 (100%) | 68 (92%) | - |  |  |
| Hepatitis B immunization | 18 (100%) | 17 (100%) | 54 (100%) | **12 (16%)** | - |  |  |
| Needle prick injury protocol | 18 (100%) | 17 (100%) | 48 (89%) | **26 (35%)** | - |  |  |
| Exposure to infectious material | 18(100%) | 17 (100%) | 52 (96%) | 58 (78%) | - |  |  |
| Hazmat exposure | 13 (72%) | 14 (82%) | 47 (87%) | **23 (31%)** | - |  |  |
| Personal protective devices used |  |  |  |  |  |  |  |
| Gloves | 18 (100%) | 17 (100%) | 54 (100%) | 68 (92%) | 63% | 94% | 100% |
| Shoe cover | 18 (100%) | 17 (100%) | 54 (100%) | 68 (92%) | **6%** | **27%** | 100% |
| Aprons /Gowns | 18 (100%) | 17 (100%) | 54 (100%) | 68 (92%) | 86% | 100% | 100% |
| Masks | 18 (100%) | 17 (100%) | 54 (100%) | 68 (92%) | 10% | 84% | 100% |
| Caps | 18 (100%) | 17 (100%) | 54 (100%) | 68 (92%) | **7%** | **16%** | 100% |
| Training received | 18 (100%) | 17 (100%) | 54 (100%) | 68 (92%) | - | - | - |
| PPE kit in hazardous places | 18 (100%) | 17 (100%) | 45 (83%) | 12 (16%) | - | **30%** | - |
| Health insurance provided | 100% corporate tie-up |  |  |  | - | - | - |

Gloves and aprons were the only protective devices used. Sweepers were not using any of the protective devices except gloves (80%). The use of protective devices was satisfactory as per the area of use. The lab workers used PPE kit in 30% incidences which in considerably low in our opinion especially in microbiology laboratory. All employees were provided health insurance by corporate tie-up with TPA (Third party administrator) and the level of coverage was as per their hierarchy and salary structure.

## Discussion

Biomedical waste management is a collective effort of many agencies and people, any breach in the process can result into failure of the whole process. The waste needs to be tackled carefully from the point of generation to the final destination keeping in mind the safety of patients, staff and environment. We conducted this study in a non-NABH accredited small healthcare organization (60 bedded) in Rajgarh city before they applied for NABH assessment in order to examine the knowledge, attitude and practices of the staff.

In our study we find that in this hospital the nurses and lab-technologists were quite knowledgeable about waste categories and their segregation and their knowledge was superior to that of housekeeping staff, although housekeeping staff did wonderful in most of categories their performance was not up to mark in some categories (viz the expired medicine), similar findings were also observed in an Indian study [13]. The good understanding is probably because of induction training and constant motivation and repetitive training of nurses and Laboratory staff by quality manger and operation manager. More than the theory it is because they were doing these practices strictly in the hospital for about 2 years. The operation and quality manager in collaboration with deputy medical superintendent and nursing superintendent were organizing the training and ward incharge nurses and nursing superintendents were responsible for observing the compliance and Nursing superintendent is responsible for holding such trainings for the new employees with the help of relevant faculty from nursing college and medical college. The senior nursing staff and sisters in charge of the ward are given the responsibility for implementation of the BMW management rules by the authorities. The housekeeping staff on the contrary did not receive repetitive training but only orientation and induction training. The housekeeping supervisor (who was a post-graduate in business administration) did receive constant training and he was responsible for training the housekeeping staff but he was already overburdened with hospital duties that he got little time for training, nevertheless the housekeeping staff performed well because they were practicing it for long and were constantly under supervision of ward-nurses. Lack of training in staff result in less knowledge in BMW management practices [14]. The hospital did not have a radiotherapy department hence the respondents were mostly blank on this aspect those who responded rightly were either had worked in some other hospital or knew it because they had studied about it out of their interest.

Table 3 depicts the knowledge and practices about segregation of the bio-medical waste right at the site of its generation. Biomedical waste must be segregated at the site of its generation to avoid appropriate disposal and to minimize health risk to the staff handling it and to the environment. The availability of color-coded bins and charts about the waste categories are important parameters which could influence the success of the BMW segregation. Because of government regulation it is a mandatory requirement for registration of a healthcare facility to comply with the BMW segregation and disposal practices. In an Indian study from Gujarat it was found that most hospitals and labs are strictly following these regulations [15]. In our observation this hospital had complied nicely with most of regulatory processes except the bar coding of bins, which was informed to be in process. The handling of sharp waste was quite impressive and ample disinfectant was available; these finding were in accordance with another Indian study from Himachal Pradesh [16].

The understanding about need of immunization was satisfactory in all the staff except the housekeeping staff where only 16% were aware of hepatitis vaccination. The hospital did not have a policy of immunization for staff but they were positive about in future. There was good awareness about needle prick injury except in housekeeping staff (35%). As far as personal protective devices are concerned there was good compliance in operation theatres. Aprons /gowns were most common equipment donned by the healthcare workers in general. The compliance with shoe-covers and caps was really poor in areas other than operation theatres. The sweepers were not given adequate protective equipment while they handle the BMW. The personal protective kits were used by laboratory workers and surgeons while handling a potentially infected patient but its availability, cost and other factors were limiting its use in 30% of cases.

The best part observed in this study was the continuous efforts, training and motivation of the healthcare workers towards proper management of BMW. There were many areas of improvement especially in training of ground work force, training about hazmat and radiological waste, and availability of personal protective equipment to everyone directly handling the potentially infected patients.

## Conclusion

Constant motivation, induction and training coupled with reinforcement in terms of feedback is vital for implementation of BMW practices. The person responsible for handling and disposal of BMW should be well informed about the potential dangers to self, others and environment. Their training should include mock drills and real-life scenarios to make thing possible in a better way.

## Data Availability

all relevant data included, any further data can be provided on request

## Author contribution

Virender Verma and Priya Soni conceptualized the study, Meenakshi Kalhan and Sanjiv Nanda helped in reviewing and statistical methods, Anjani Kumar and Vivek Nandan helped in designing of questionnaires, data management, and drafting.

## References

1. Padmanabhan KK, Barik D. Health Hazards of Medical Waste and its Disposal. Energy from Toxic Organic Waste for Heat and Power Generation. 2019:99–118. doi: https://doi.org/10.1016/B978-0-08-102528-4.00008-0.

2. Stern PC, Young OR, Druckman D. Human Causes of Global Change. In: Stern PC, Young OR, Druckman D, editors. GLOBAL ENVIRONMENTAL CHANGE: Understanding the Human Dimensions. Washington D.C, USA: National Research Council, NATIONAL ACADEMY PRESS 1992. p. 44–99.

3. W.H.O: Health-care waste. https://www.who.int/news-room/fact-sheets/detail/health-care-waste (2018). Accessed 15-may 2020.

4. Indian-governent: BIO-MEDICAL WASTE (MANAGEMENT AND HANDLING) RULES, 1998.https://hspcb.gov.in/BMW%20Rules.pdf (1998). Accessed 15-may 2020.

5. Mandal SK, Dutta J. Integrated Bio-Medical Waste Management Plan for Patna City. Institute of Town Planners, India Journal. 2009;6(April June 2009):1–25.

6. Shahab S, Hashmi S. Hospital and biomedical waste management. In: M I, editor. Community Medicine and Public Health.Karachi: Time Publishers; 2003. p. 426–37.

7. Horvath A. Management of waste disposal in medical institutions. Rv Hetil. 1991;192:919–24.

8. Chandrorkar A, Nagoba B. Hospital Waste Management. 1 ed. Hyderabad: Paras Medical Publisher; 2003; 2003.

9. Government-of-India: Hospitals in the Country. https://pib.gov.in/PressReleasePage.aspx?PRID=1539877 (2018). Accessed 15-may 2020.

10. Hendricks R, Pool EJ. The effectiveness of sewage treatment processes to remove faecal pathogens and antibiotic residues. J Environ Sci Health A Tox Hazard Subst Environ Eng. 2012;47(2):289–97. doi: 10.1080/10934529.2012.637432.

11. Author: Robert D Cox ZFD: Hazmat. https://emedicine.medscape.com/article/764812-overview (2018). Accessed 15-may 2020.

12. Robert D Cox ZFD: Hazmat. https://emedicine.medscape.com/article/764812-overview (2018). Accessed 15-may 2020.

13. Saraf Y, Shinde M, Tiwari S. Study of awareness status about hospital waste management among personnel and quantification. Indian J Community Med. 2006;31(111).

14. Soliman SM, Ahmed AL. Overview of biomedical waste management in selected governorates in Egypt: a pilot study. Waste Management. Waste management. 2007:1920–3.

15. Pandit NB, Mehta HK, Kartha GP. Management of biomedical waste: awareness and practices in a district of Gujarat. Ind J Pub Health. 2005;49:245–7.

16. Kumar S, Mazta SR, Gupta AK, 3. Comparing the Biomedical Waste Management Practices in Major Public and Private SectorHospitals of Shimla City. International Journal of Scientific Study. February 2015;2(11).

